# Microvascular blood flow changes of the abductor pollicis brevi muscle during sustained static exercise

**DOI:** 10.1101/2020.06.03.20120931

**Authors:** Martina Giovannella, Evelina Urtane, Umut Karadeniz, Uldis Rubins, Udo M. Weigel, Zbignevs Marcinkevics, Turgut Durduran

## Abstract

A practical assessment of the health of the palm muscle, abductor pollicis brevis (APB), is important for diagnosis of different conditions. Here we have developed a protocol and a probe to utilize diffuse correlation spectroscopy (DCS) to characterize microvascular blood flow changes in the APB during and after sustained isometric exercise, during and after thumb abduction at at 55% of maximum voluntary contraction (MVC). Blood flow in the APB decreased during exercise in the subjects (n=13) with high MVC (n=7) and stayed constant in the ones with low MVC (n=6) suggesting that the mechanical occlusion due to increased intramuscular pressure exceeded the vasodilation elicited by the higher demand. Blood flow changes during exercise negatively correlated with the absolute force applied by each subject. Muscular blood flow increased after exercise compared to the values reached during exercise. In conclusion, DCS allows the study of the response of a small muscle to static exercise.

## 1. INTRODUCTION

The metabolic demand of skeletal muscles increases during exercise and the assurance of proper blood perfusion is fundamental for the muscle well-being. However, t it canbe compromised in various pathological conditions [1–4]. In particular, small palmar muscles are often compromised by daily manipulations such as heavy computer usage and extensive manipulation of a hand-held device [5]. The continuous overuse of small palm muscles that control fine movements of the thumb (thenar muscles) creates damage and inflammation resulting in the stiffening and pain in the thenar eminence (soft muscular rounded part at the base of the thumb) causing the so called repetitive strain injury. Of particular interest is the assessment of blood perfusion to this area as an indicator of proper thenar muscle function.

This work focuses on the abductor pollicis brevis (APB) among the thenar muscles. This is the most lateral and superficial of the three muscles forming the thenar eminence and it is a flat, thin, triangular fusiform muscle. It is the smallest intrinsic thenar muscle and one of the few intrinsic muscles innervated by the median nerve with the large number of small motor units [6,7]. APB’s main function is to move the thumb away from the palm in a perpendicular direction of the surface of the palm (palm abduction) [8–10] and it is essential for hand functioning [11]. Its functions can be compromised by the carpal tunnel syndrome (CTS) [12] which can cause it to weaken and atrophy [13]. Interestingly, the assessment of the atrophy and the strength of the APB during exercise can useful for CTS diagnosis [14] and can predict the failure or success of surgical treatment [15]. In addition, vascular changes have been proven to precede electrophysiological abnormalities in patients with evidence of CTS [16]. However, up to now, the blood perfusion to the thenar muscles has only been monitored by means of Doppler ultrasonography in the forearm [17, 18], which provides information on the large vessels that irrigate more than one muscle. Better, practical methods are still needed to evaluate local, specific biomarkers of impaired blood perfusion.

Beyond Doppler ultrasonography, various methods have been explored to measure blood perfusion in exercising skeletal muscle [4] but none of them are satisfactory for measuring the local muscle blood flow. Among the cost-effective, non invasive, continuous and easily available options laser Doppler [19] and venous occlusion plethysmography [20] are promising. Nonetheless, the former is restricted to shallow (< 1 mm) tissue measurements, while the latter requires the interruption of venous blood flow, limiting the protocols it can be applied in.

Near-infrared spectroscopy technologies are good candidates for monitoring the haemodynamics of skeletal muscle thanks to their non-invasiveness and cost-effectiveness [21]. Typical near-infrared spectroscopy (NIRS) measures microvascular blood oxygen saturation dynamics [22, 23]. Among those, diffuse correlation spectroscopy (DCS) [24-26] uses near infrared (NIR) laser light to measure microvascular blood flow in the microvasculature of the deep (~1 cm) tissue. DCS has already been validated for muscle blood flow measurement [27] and was used in various protocols to measure perfusion of large skeletal muscles, for example on the thigh, the leg and the forearm mainly during dynamic exercise protocols [28].

The aim of this study is to assess blood flow in APB during sustained static exercise of moderate intensity and during post-exercise recovery by means of DCS. We chose to perform moderate intensity exercise in order to minimize the systemic response to the exercise which depends on the muscle volume and exercise intensity [29,30]. Therefore, the results here presented reflect the local perfusion response to the exercise.

## 2. METHODS

### 2.1. Diffuse optical monitors for muscle haemodynamics and data analysis

Two different diffuse optical monitors were employed in this study. Firstly, time resolved near infrared spectroscopy (TRS) was used to measure the APB optical properties. TRS measures the broadening of a few hundreds of picoseconds wide laser pulse after its propagation through the tissue. By modeling the shape of the collected pulse, absorption (μ_a_) and reduced scattering coefficient (μ’_s_) at the wavelengths used are obtained [31]. Specifically, a prototype of a TRS device (TRS-20, Hamamatsu Photonics K.K., Japan) was used. This device has two source-detector channels and has three laser sources at 760 nm, 800 nm and 830 nm. Only one of the two channels was used. The separation between source and detection fiber was 15 mm with the fibers embedded in a black foam probe. The collected distribution of time-of-flight of photons measured by TRS was fitted to the analytical solution for the reflectance in a semi-infinite homogeneous medium [32, 33] in order to retrieve the optical properties of the tissue probed.

Diffuse correlation spectroscopy (DCS) was used to obtain a measurement of the blood flow in the microvasculature of the muscle. DCS measures the intensity fluctuations of a single speckle resulting from the propagation of NIR long coherence length laser light through the tissue. Those intensity fluctuations are affected by the movement of the scatterers present in the tissue, dominated by the red blood cells [24]. By studying the decay of the intensity autocorrelation function a measure of the diffusion coefficient of moving scatterers can be retrieved [25]. This quantity, usually referred to as blood flow index (BFI) has units of cm^2^/s and is proportional to the blood flow [26, 34]. A commercial DCS device (HemoFloMo, HemoPhotonics S.L., Spain) was used in this experiment. It uses a long coherence length 785 nm laser and it has four parallel source channels and eight parallel detector channels. The probe employed one of the source and two of the detector channels. One detection fiber was placed at 8 mm from the source, while the other at 15 mm. All three were embedded in a black foam probe. DCS measurements were analyzed using the solution for the diffusion equation of the electric field autocorrelation function for the semi-infinite medium [25, 26] to derive the BFI. This analytical function contains the optical properties of the tissue, which must be introduced as input parameters [35]. Optical properties measured by TRS at 800 nm were used for the DCS analysis.

### 2.2. Protocol

Healthy, right-handed adult subjects were recruited for a single measurement session. The study was approved by the ethical committee of Hospital Clinic Barcelona (Spain). Each subject signed an informed consent and the study was conducted according to the principles of the Declaration of Helsinki.

To start with, the subject was seated on a comfortable reclined chair with the right hand lying on a vacuum pillow. This was adapted to the hand shape to avoid movement and to keep the hand at the heart level. The area of the APB muscle was inspected by an ultrasound in B mode (Sonosite Titan L38 transducer, UMI, US) and an image was acquired for each subject in order to calculate the thickness of skin, adipose tissue and muscle layer. These measurements were carried out to investigate the role of the adipose tissue thickness as a confounding factor but not for the use of a layered tissue model for data analysis which is beyond the scope of this work.

The TRS probe was placed on the same area to acquire 30 s of measurement to estimate the optical properties. Afterwards the DCS probe was attached approximately on the same spot. A force transducer (Flexi Forxe Sensor III, Tekscan, US) was kept on the external part of the thumb by a non-elastic band placed over the hand to avoid movements of the thumb. The force measured as a voltage by the force sensor was calibrated by pressing the sensor over a scale resulting in a conversion factor of 1.3 kg/V. A screen (Gauge Indicator, LabVIEW, US) displayed the reading of the force sensor on-line. Once the DCS probe, the force transducer and the band were appropriately placed (figure 1), the maximum voluntary contraction (MVC) was measured. To do so, the subject was asked to press the thumb against the belt to measure the maximum force maintained for at least 5 s. The process was done three times and the average recording used as the MVC.

**Fig. 1.**
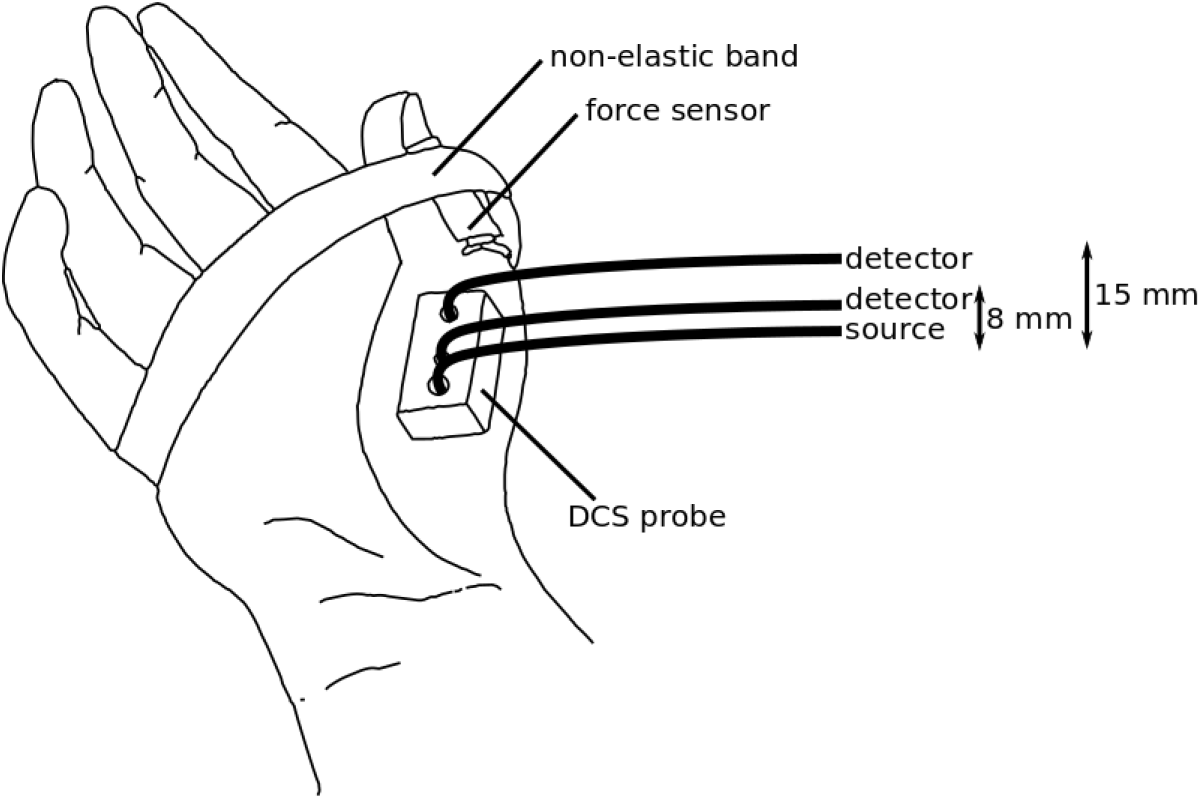
The placement of the DCS probe on the APB muscle. One detector fiber was kept at a distance of 8 mm, while the other was at 15 mm from the source fiber. The thumb was kept attached to the hand by a non-elastic band, which held a force transducer in order to measure the strength applied toward the band by the thumb. Subject’s hands were placed on a vacuum pillow support to avoid movement (not shown in the figure).

At this point, DCS acquisition was started. After three minutes of rest, the subject was asked to press the thumb towards the belt applying 55% of the MVC and maintain its pressure constant for 30 s following the force sensor recording displayed on the screen. Afterwards the subject rested for three minutes and another thirty seconds of exercise at 55% of the MVC followed. Analogously, three minutes of rest were measured after the second exercise. DCS measurements were acquired at 1 s sampling time, but the acquisition was paused for 5 s every five acquired measurements, in order to record acquisitions by another device which is not related to the goal of this study.

The mean arterial pressure (MAP), the heart rate (HR) and the cardiac output (CO) were measured over the whole protocol by a non invasive continuous blood pressure monitor (Finometer Midi, Finapres, Netherlands) applied over the non-exercising hand.

### 2.3. Statistical data analysis

To start with, median, first and third interquartile values were calculated for layer thicknesses, MVC, optical properties, baseline BFI and baseline physiological parameters. Here, the baseline period was defined as the 60 s before exercise. Subsequently, the change with respect to the baseline was calculated for all the physiological parameters measured by the Finapres (∆MAP, ∆HR, ∆CO). On the other hand, BFI was normalized to the baseline to derive a relative BFI (rBFI) and a rBFI percentage change (∆rBFI). It must be noted that the measurements that were acquired by DCS right before and right after the exercise was excluded since they may be influenced by motion artifacts. The calculation of changes with respect to the baseline was done independently for the two exercise repetitions.

In order to characterize the response to exercise, the change of physiological parameters and rBFI was evaluated for each subject in four time windows. The onset of the exercise was marked as t=0 s then the following time windows were defined; (1) during exercise (“During”) from t=15 s to t=30 s; (2) right after exercise (“Post1”) at t=40±5 s; (3) “Post2” as t=80±5 s, and, (4) “Post3” at t=160±5 s.

Since the goal was to study the local response of microvascular, blood perfusion in the muscle to the exercise, and to avoid contamination of the physiology by systemic changes, we have tested whether CO, MAP and HR changed during and after exercise. All the three parameters were evaluated at each of the four data points explained above for the two exercise repetitions by using a Wilcoxon signed-rank test.

Finally, the response of microvascular blood flow to exercise was evaluated. First, we have tested whether the two exercise repetitions triggered a different response. To do so, a linear mixed effect (LME) model was built with the exercise repetition as the fixed effect and the subject identifier as the random effect for each of the four selected time windows and source detector separations.

The subjects were divided in two groups considering the median of the MVC. The ones with lower MVC than the median were included in the low MVC group, while the others in the high MVC group.

Wilcoxon signed-rank test was used to test whether rBFI changed respect to baseline in each of the four time windows. In addition a paired Wilcoxon signed-rank test was run between During and Post1 time points, in order to test whether there is a further change after the end of the exercise.

To conclude with, we have aimed to check whether the change in blood flow during and after exercise depends on the absolute force applied by each subject, whose MVC is a surrogate measure. Therefore, a correlation between ∆rBFI in the four time windows and the MVC was tested through calculation of the Pearson coefficient (R). Statistical data analysis was performed through R [36] and threshold for significance for p-values was defined as 0.05 for all the statistical tests.

## 3. RESULTS

Thirteen subjects (n=13) were recruited for the study. An example of the ultrasound image is shown in figure 2(a). The thicknesses of the skin, adipose subcutaneous tissue and muscle layer are shown in figure 2(b) for each subject. The median of the superficial layer thickness (skin and adipose tissue layer) was 2.1 mm (1.7 mm, 2.4 mm). The first and third interquartile are shown in brackets throughout the rest of the text. The median thickness of APB muscle was 6.3 mm (5.2 mm, 7.6 mm).

**Fig. 2.**
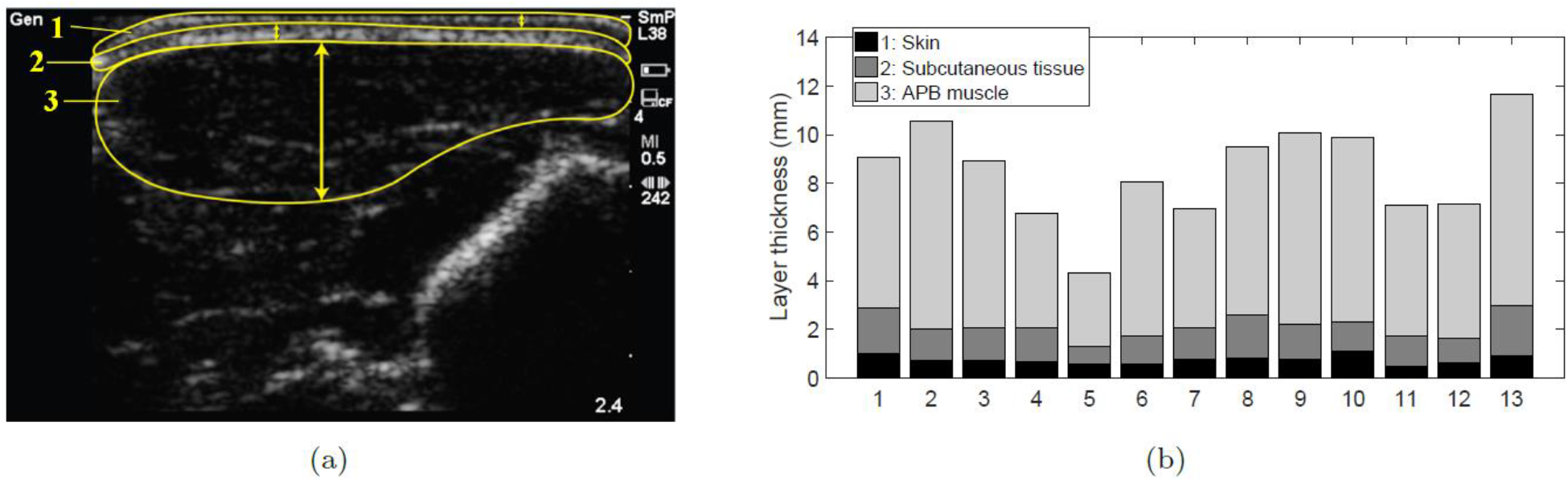
**(a)** An ultrasound image acquired from one subject. Three different structures can be defined: the skin (1), the adipose subcutaneous tissue (2) and the abductor pollicis brevis (APB) muscle (3). **(b)** The three layer thicknesses measured in all the subjects. The skin and adipose subcutaneous tissue will be referred as superficial layer for the rest of the manuscript.

The MVC measured for each subject is reported in figure 3 and the median is 2.3 kg (1.9 kg, 2.9 kg). The baseline values for physiological parameters, optical properties and BFI are summarized in table 1. Furthermore, the optical properties and BFI are shown for each subject in figure 4.

**Fig. 3.**
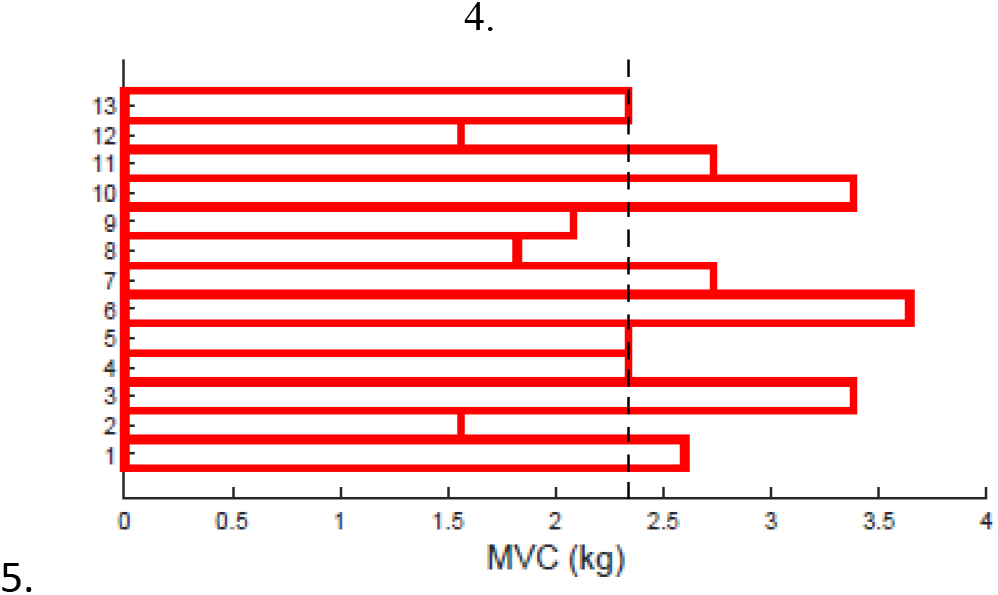
MVC measured in the 13 subjects. Median (first, third interquartile) is 2.3 kg (1.9 kg, 2.9 kg). The black vertical line highlights the separation between high (N=6) and low (N=7) MVC group.

**Fig. 4.**
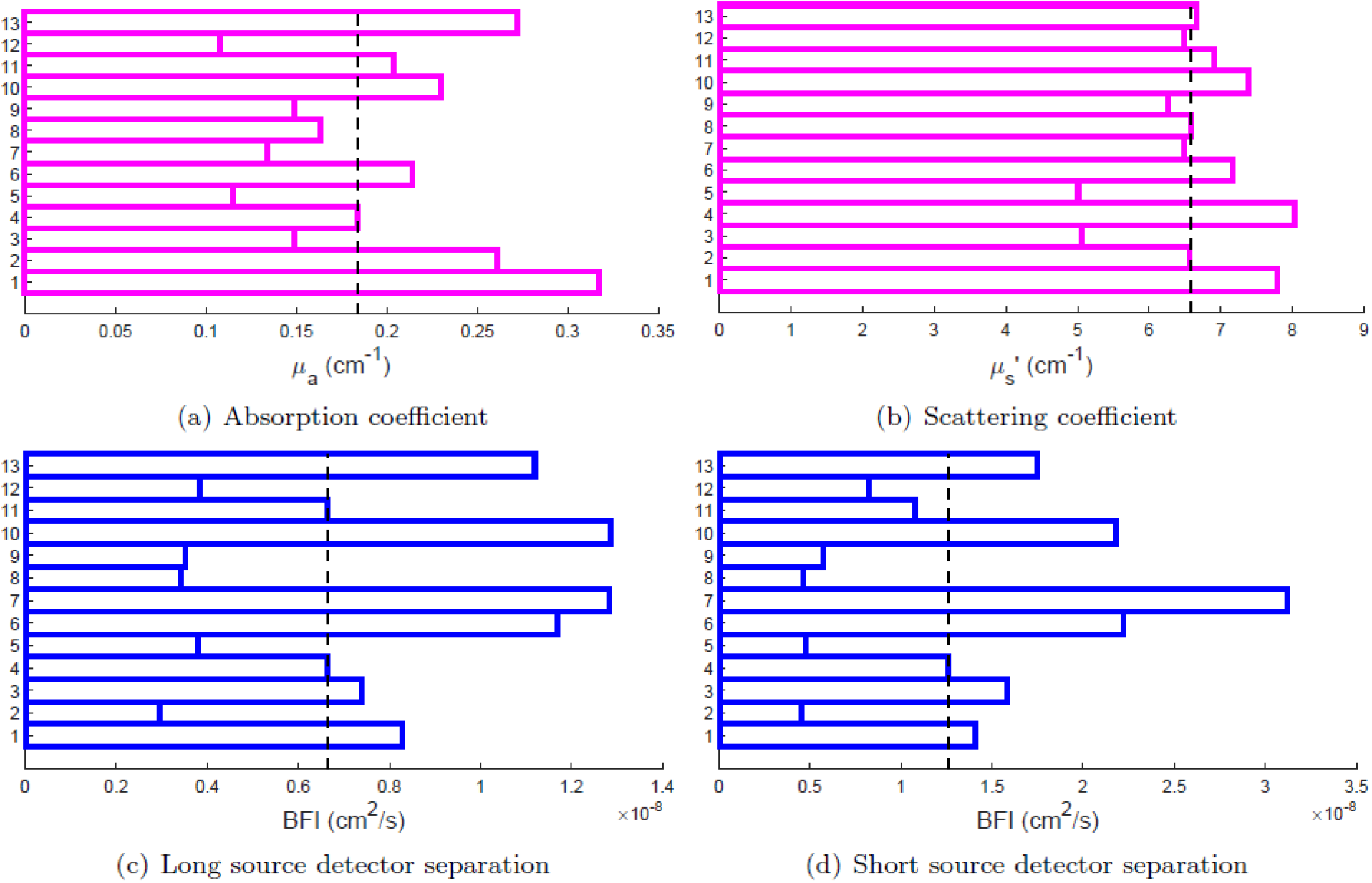
**(a) and (b)**: Optical properties at 800 nm measured in the 13 subjects by the TRS, absorption and scattering coefficient, respectively. (c) and (d): BFI measured in the 13 subjects by DCS, from the long and short source detector separation, respectively. The dashed vertical line represents the median, first and third interquartile are reported in table 1.

**Table 1.** Median (first, third interquartile) of baseline values over all the subjects. Mean arterial pressure (MAP), heart rate (HR), cardiac output (CO), absorption (μ_α_) and reduced scattering coefficient (μ’s) at 800 nm and blood flow index (BFI) as measured by the two source detector separations are reported.

| Physiological parameters |  |
| --- | --- |
| MAP | 88 (84,98) mmHg |
| HR | 67 (62,77) bpm |
| CO | 7.0 (5.3,8.6) L/min |
| Optical properties @800 nm |  |
| $\mu_a$ | 0.18 (0.14, 0.24) $\text{cm}^{-1}$ |
| $\mu'_s$ | 6.6 (6.4, 7.22) $\text{cm}^{-1}$ |
| Blood flow index (BFI) |  |
| Long separation (15 mm) | $0.7 (0.4, 1.4) \times 10^{-8} \text{ cm}^2/\text{s}$ |
| Short separation (8 mm) | $1.0 (0.6, 2.2) \times 10^{-8} \text{ cm}^2/\text{s}$ |

As argued before, the first step was to check whether the protocol causes changes in the systemic circulation. MAP did not change during the whole protocol, while a small increase was registered for HR and CO only during the second exercise. In particular, HR increased by 3.5 bpm (5% of the baseline, p=0.007) and CO by 0.5 L/min (7% of the baseline, p=0.01).

LME analysis (see above) confirmed that the BFI response was equivalent in the two exercise repetitions throughout the four time windows (p>>0.05 for all cases).

Figure 5 shows the average time series of ΔrBFI for the two groups (high and low MVC) and the two DCS source detector separations. Figure 6 shows traditional boxplots of ∆rBFI during the four time windows of interest. Red stars highlight the statistically significant difference from zero (p<0.05) as tested by the Wilcoxon signed-rank test. During exercise a decrease in BFI was detected in the high MVC group, while it stayed constant for the low MVC group. Triangles in figure 6 highlight statistically significant changes (p<0.05) of rBFI after the exercise with respect to the values reached during exercise. After exercise, blood flow increased in the low MVC group as registered by both long and short source detector separations, while in the high MVC group statistical significance was observed only by the long source detector separation. The increase in blood flow reached a higher level than the baseline in the low MVC group and it did not recover even 80 s after the exercise. It recovered by 160 s for both for short and long source detector separations.

**Fig. 5.**
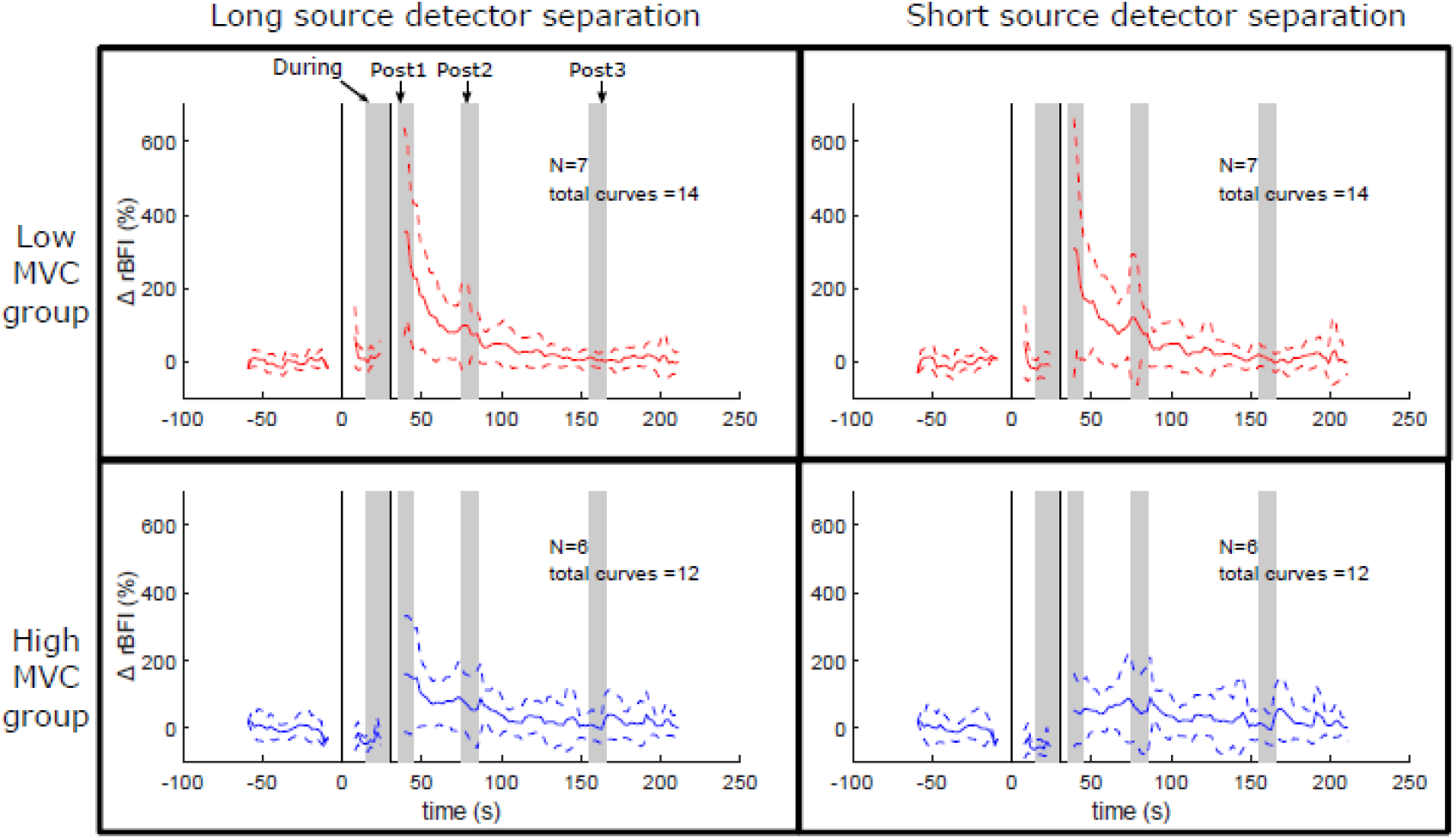
APB muscle blood flow change during and after 30 s of sustained thumb abduction at 55% MVC. The onset of the exercise is a time t=0 s and the vertical black lines highlight the exercise period. Results from the group with low MVC (N=7) are shown in red in the upper row, while results from the group with high MVC (N=6) in blue in the bottom one. The long separation is displayed in the left column and the short in the right one. The four time windows of interest are highlighted by grey vertical bars.

**Fig. 6.**
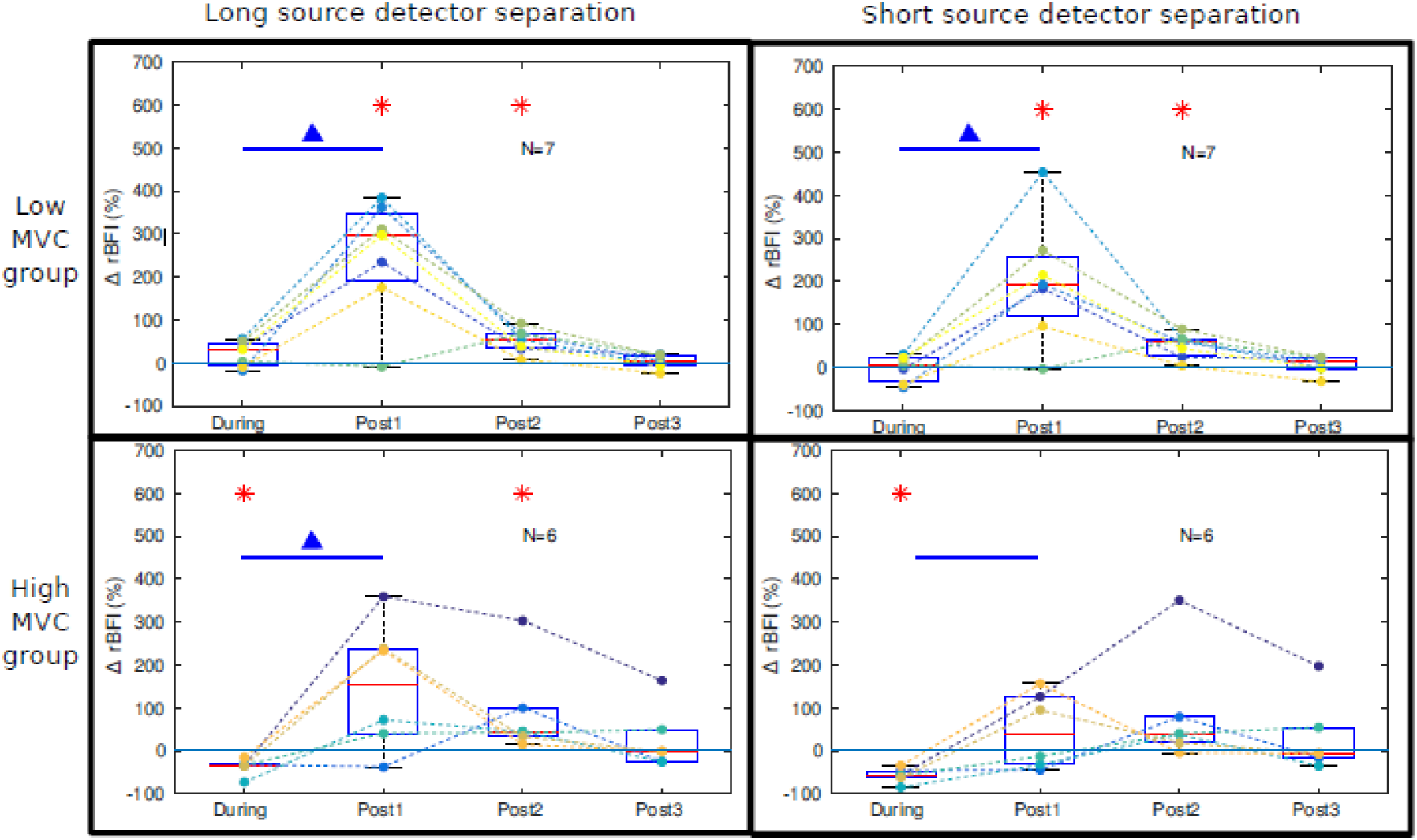
Boxplot of ∆rBFI for the two source-detector separations (columns) divided according to the MVC group (rows). The four periods correspond to “During” from 15 s to 30 s, i.e. during exercise (start at 0 s) and post-exercise “Post1” at 40 s, “Post2” at 80 s and “Post3” is at 160 s. Red stars indicate statistically significant difference from zero. Blue triangles indicate statistically significant difference between “During” and the tested period. Please see text for further details.

Lastly, the correlation between the ∆rBFI in each of the four time windows considered and the MVC measured in each subject was tested. A statistically significant correlation was found only in the period during the exercise both for the long (R=-0.7, p=0.009) and the short source detector separation (R=-0.6, p=0.02). Others were non-significant.

## 4. DISCUSSION

This work is, to the best of our knowledge, the first to measure alterations of local blood flow in the abductor pollicis brevis (APB) muscle in response to exercise by diffuse correlation spectroscopy (DCS). APB was selected since is involved in important hand functions and its performance can be compromised by musculo-skeletal disorders of the upper extremities. In addition, it is an interesting muscle to consider as a model to study the microvascular blood flow dynamics that are affected by the local physiology rather than a larger reaction that alters the systemic physiology. This was hypothesized because a moderate intensity of exercise of a small muscle like APB, whose total blood supply is rather limited [7] is not expected to influence the systemic circulation [29]. Our results indicate this to be true for the first period of exercise with a minimal, possibly stress induced change in the systemic physiology during the second repetition. Overall, our findings are within physiological expectations and illustrate that DCS is indeed a potential technology to further explore and adapt for understanding local muscle physiology. We now discuss specific results and limitations of the study.

The protocol and the subject population was verified to be appropriate since an indicator, MVC in each subject (figure 3) was found to be comparable to previously published results [10, 37, 38]. In order to verify how the protocol affected systemic circulation, the heart rate (HR), mean arterial blood pressure (MAP) and cardiac output (CO) were monitored in the protocol. This recording showed that MAP did not change along the whole protocol, while we have found a small increase in the second exercise for HR and CO of 3 bpm and 0.5 L/min respectively. Different reasons may have lead to this increase in the second repetition of exercise. Subject may have been more stressed during the second repetition with respect to the first one or they may have involved more muscles because they knew what to expect. The measured changes are far smaller than what was reported for for moderate intensity exercises of larger muscles [30, 39] and we do not expect this to influence the muscle blood flow. Furthermore, the blood flow response as measured by DCS did not differ between the two exercise repetitions.

Perfusion response to sustained isometric exercise is currently not fully understood because of different factors playing roles in the process. On one hand, the metabolic demand of the muscle is higher and this leads to the release of metabolites to ensure vasodilation in the interested muscle [40]. On the other hand, the contraction increases the intramuscular pressure and mechanically occludes the vessels in that region, partially or totally. The pressure depends on the contraction force and it has been observed that moderate intensity exercises (50% of MVC) lead to a decreased or unchanged perfusion to the muscle in the first tens of seconds of exercise [41–43]. This is different from dynamic rhythmic exercise during which muscle perfusion increases due to the relaxation period [44]. Along with these observations in literature, overall, the blood flow as measured by DCS in our experiment does not increase during static exercise.

Interestingly, we have observed a different behavior in subjects with high MVC, who performed exercise at a stronger force, compared to the low MVC ones. In particular, muscle blood flow decreased during exercise in the high MVC group, while it stayed constant in the low MVC group. This suggests that the intramuscular pressure depends not only on the relative intensity with respect to the MVC but also on the absolute force which is higher in the high MVC group. This type of relationship has been discussed in literature [42, 45]. We have indeed found a statistically significant negative correlation between the change of blood flow during exercise and the MVC measured in each subject. The higher the MVC, *i.e*. the force applied, the lower the blood flow during exercise, due to the increase intramuscular pressure. This topic can be further explored using different protocols and in a larger population.

After the exercise, the blood flow increased with respect to the exercise period and, for the low MVC group, it reached higher levels than baseline. This was expected due to the previously elicited vasodilation since muscle relaxation stops the mechanical occlusion.

Comparable, but not equivalent, results were registered by the long and the short source detector separations. It must be reminded that DCS measures a mixture of extra-and intra-muscular perfusion and the weight of the former is more prominent in the short source detector separation. These results confirm that, even if the superficial layer on top of the APB is very thin, it partially masks the muscle perfusion to the detector placed at 8 mm from the source. We note that it was feasible to use a probe with an intra-fiber distance of 15 mm both in terms of the muscle geometry and the signal intensity. This finding may be relevant for other applications that rely on thenar hemodynamics in the intensive care units such as those that utilize hemodynamics to evaluate for sepsis [46] for weaning from ventilators [47]. In these applications, thenar muscle is utilized since it has less anatomical variability between subjects and is accessible.

DCS continuously and non-invasively assesses the local blood flow in relatively deep tissues. Being cost-effective and portable, it can be applied to a variety of protocols and conditions. However, DCS is sensitive to fiber and probe motion. For this reason its usage in dynamic, rhythmically exercise requires signal gating in order to exclude the time periods corresponding to limb movement [48–50]. We have chosen to perform an isometric exercise in order to avoid motion artifacts in DCS recording. This avoids the complications of monitoring movements and makes the experiment more feasible and easily reproducible, enlarging the possibility of applications.

## 5. CONCLUSION

We have described the microvascular blood flow response in the APB during and after moderate intensity static exercise in healthy subjects. This was made possible by employing DCS, a non-invasive optical technology that allows us to continuously assess the microvascular blood flow in the muscle tissue. Since the global systemic response to exercise stress is minimized in this kind of exercise, we have monitored the local mechanism that regulates the blood perfusion to the oxygen demanding muscle. Blood flow was decreased or unchanged during 30 s of sustained isometric exercise depending on the absolute force applied. This suggested that mechanical occlusion due to increased intramuscular pressure exceeded the vasodilation consequent to the higher demand of oxygen during contraction. When the muscle was relaxed, the occlusion was opened and an increase of blood flow respect to the exercise period (and respect to baseline for low MVC group) was registered.

## Data Availability

The data that support the findings of this study are available from the corresponding author, [M Giovannella], upon reasonable request.

## ACKNOWLEDGEMENTS & FUNDING

This research was funded by Fundació CELLEX Barcelona, Ministerio de Economía y Competitividad / FEDER (PHOTODEMENTIA, DPI2015-64358-C2-1-R), Instituto de Salud CarlosIII / FEDER (MEDPHOTAGE, DTS16/00087), the “Severo Ochoa” Programme for Centres of Excellence in R&D (SEV-2015-0522), the Obra social “la Caixa” Foundation (LlumMedBcn), Institució CERCA, AGAUR-Generalitat (2017 SGR 1380), LASERLAB-EUROPE IV (GA: 654148) and European Regional Development Fund project 2014/0036/2DP/2.1.1.1.0/14/APIA/VIAA/020. Martina Giovannella’s Ph.D. was funded by the “Severo Ochoa” Programme for Centres of Excellence in R&D (SEV-2015-0522). We thank Hamamatsu Photonics K.K. and its Spain office for loaning the TRS-20 system and for their collaboration.

